# Hidden periods, duration and final size of COVID-19 pandemic

**DOI:** 10.1101/2020.05.10.20097147

**Authors:** Igor Nesteruk

## Abstract

The SIR (susceptible-infected-removed) model, statistical approach for the parameter identification and the official WHO data about the confirmed cumulative number of cases were used to estimate the characteristics of COVID-19 pandemic in USA, Germany, UK, South Korea and in the world. Epidemic in every country has rather long hidden period before fist cases were confirmed. In particular, the pandemic began in China no later than October, 2019. If current trends continue, the end of the pandemic should be expected no earlier than March 2021, the global number of cases will exceed 5 million.

## Introduction

Here we consider the global COVID-19 pandemic dynamics and epidemic outbreaks in USA, Germany, UK, South Korea other countries and regions with the use of official WHO data sets, [1]. The SIR model, connecting the number of susceptible *S*, infected and spreading the infection *I* and removed *R* persons, was applied [2-4]. The unknown parameters of this model can be estimated with the use of the cumulative number of cases *V=I+R* and the statistics-based method of parameter identification developed in [5, 6].

This approach was used in [6-14] to estimate the Corona pandemic dynamics in China, the Republic of Korea, Italy, Austria, Spain, Germany, France, the Republic of Moldova, Ukraine and Kyiv. Usually the number of cases registered during the initial period of an epidemic is not reliable, since many infected persons are not detected. That is why the correct estimations of epidemic parameters can be done with the use of data sets obtained for later periods of the epidemic when the number of detected cases is closer to the real one. This fact necessitates a periodic reassessment of the epidemic’s characteristics and forecasts for its final size and duration. In this paper we will recalculate the pandemic parameters for Germany and South Korea, provide estimations UK, USA and world. The accuracy of the method and the problem of hidden periods of the epidemic outbreaks in different countries will be discussed. Some recommendation about the quarantine mitigation will be proposed.

## Materials and Methods

### Data

The official information about the accumulated numbers of confirmed COVID-19 cases *V_j_* in South Korea, Germany, UK, USA and in the world from WHO daily situation reports (numbers 81-109), [1] is presented in Table 1. The corresponding moments of time *t_j_* (measured in days) are also shown in this table. Data sets for the period April 9-29 were used for calculations. Other values were used only for verifications of calculations.

**Table 1.**
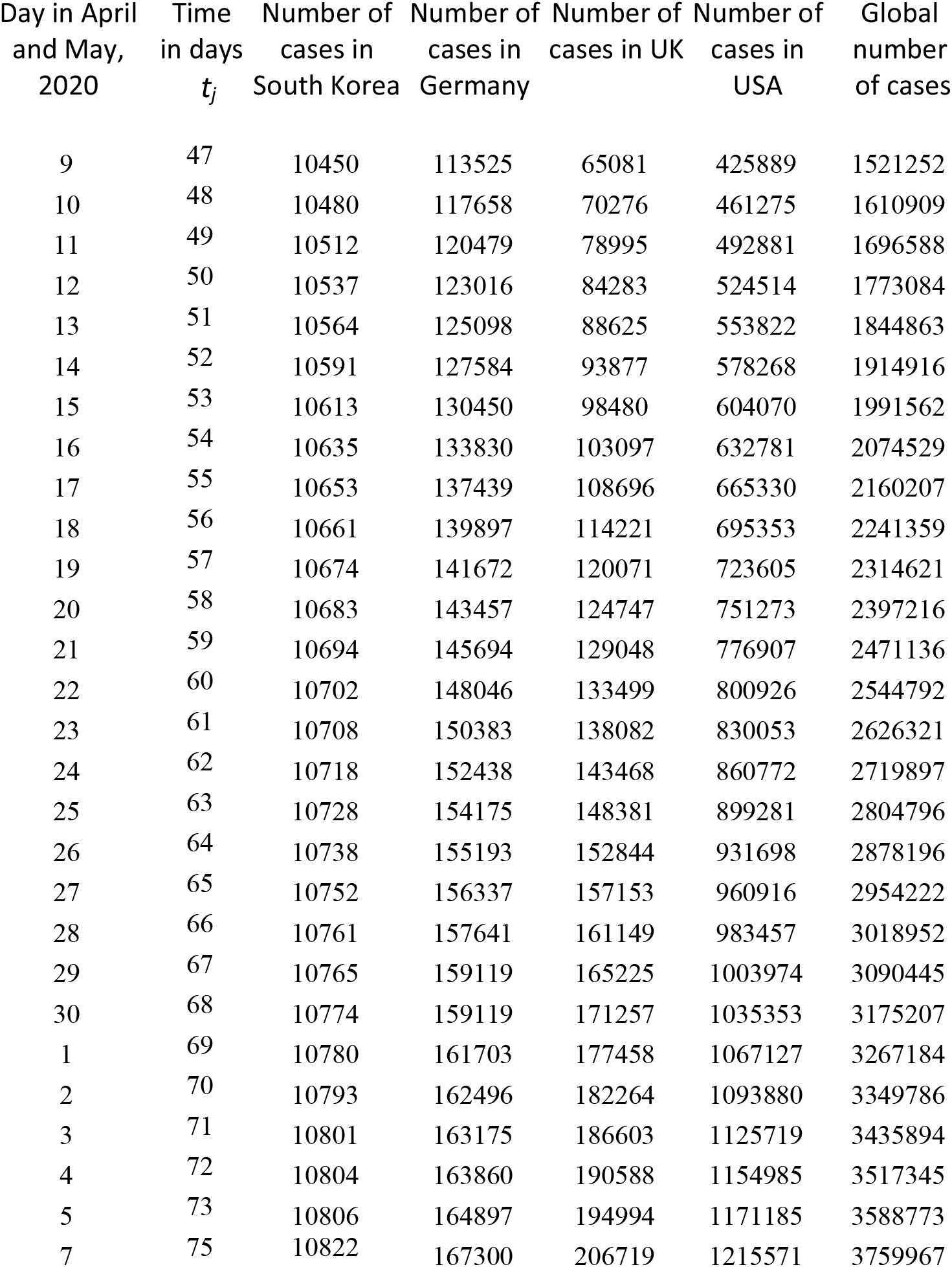
Official cumulative numbers of confirmed cases in the Republic of Korea, Germany, UK, USA and in the world used for calculations and verifications of predictions, [1].

| Day in April<br>and May,<br>2020 | Time<br>in days<br>$t_j$ | Number of<br>cases in<br>South Korea | Number of<br>cases in<br>Germany | Number of<br>cases in UK | Number of<br>cases in<br>USA | Global<br>number<br>of cases |
| --- | --- | --- | --- | --- | --- | --- |
| 9 | 47 | 10450 | 113525 | 65081 | 425889 | 1521252 |
| 10 | 48 | 10480 | 117658 | 70276 | 461275 | 1610909 |
| 11 | 49 | 10512 | 120479 | 78995 | 492881 | 1696588 |
| 12 | 50 | 10537 | 123016 | 84283 | 524514 | 1773084 |
| 13 | 51 | 10564 | 125098 | 88625 | 553822 | 1844863 |
| 14 | 52 | 10591 | 127584 | 93877 | 578268 | 1914916 |
| 15 | 53 | 10613 | 130450 | 98480 | 604070 | 1991562 |
| 16 | 54 | 10635 | 133830 | 103097 | 632781 | 2074529 |
| 17 | 55 | 10653 | 137439 | 108696 | 665330 | 2160207 |
| 18 | 56 | 10661 | 139897 | 114221 | 695353 | 2241359 |
| 19 | 57 | 10674 | 141672 | 120071 | 723605 | 2314621 |
| 20 | 58 | 10683 | 143457 | 124747 | 751273 | 2397216 |
| 21 | 59 | 10694 | 145694 | 129048 | 776907 | 2471136 |
| 22 | 60 | 10702 | 148046 | 133499 | 800926 | 2544792 |
| 23 | 61 | 10708 | 150383 | 138082 | 830053 | 2626321 |
| 24 | 62 | 10718 | 152438 | 143468 | 860772 | 2719897 |
| 25 | 63 | 10728 | 154175 | 148381 | 899281 | 2804796 |
| 26 | 64 | 10738 | 155193 | 152844 | 931698 | 2878196 |
| 27 | 65 | 10752 | 156337 | 157153 | 960916 | 2954222 |
| 28 | 66 | 10761 | 157641 | 161149 | 983457 | 3018952 |
| 29 | 67 | 10765 | 159119 | 165225 | 1003974 | 3090445 |
| 30 | 68 | 10774 | 159119 | 171257 | 1035353 | 3175207 |
| 1 | 69 | 10780 | 161703 | 177458 | 1067127 | 3267184 |
| 2 | 70 | 10793 | 162496 | 182264 | 1093880 | 3349786 |
| 3 | 71 | 10801 | 163175 | 186603 | 1125719 | 3435894 |
| 4 | 72 | 10804 | 163860 | 190588 | 1154985 | 3517345 |
| 5 | 73 | 10806 | 164897 | 194994 | 1171185 | 3588773 |
| 7 | 75 | 10822 | 167300 | 206719 | 1215571 | 3759967 |

### SIR model

The SIR model for an infectious disease [2-5] relates the number of susceptible persons *S* (persons who are sensitive to the pathogen and **not protected**); the number of infected is *I* (persons who are sick and **spread the infection**; please don’t confuse with the number of still ill persons, so known active cases) and the number of removed *R* (persons who **no longer spread the infection**; this number is the sum of isolated, recovered, dead, and infected people who left the region); *α* and *ρ* are constants.

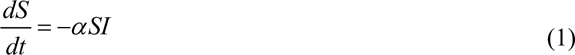

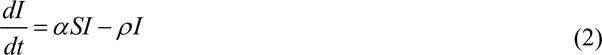

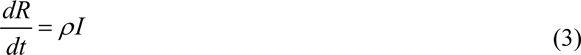

To determine the initial conditions for the set of equations (1–3), let us suppose that at the moment of the epidemic outbreak *t*_0_, [5, 6]:

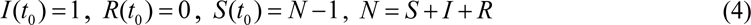

The analytical solution for the set of equations (1–3) was obtained by introducing the function *V* (*t*) = *I* (*t*) + *R*(*t*), corresponding to the number of victims or cumulative confirmed number of cases, [5, 6]:

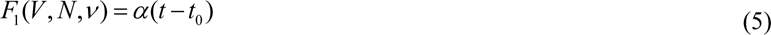

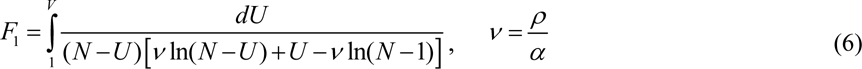

Thus, for every set of parameters *N*, *ν*, *α*, *t*_0_ and a fixed value of *V* the integral (6) can be calculated and the corresponding moment of time can be determined from (5). Then functions *I*(*t)* and *R*(*t)* can be easily calculated with the of formulas, [5, 6].

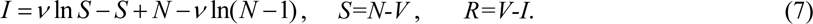

Function *I* has a maximum at *S = ν* and tends to zero at infinity, see [2, 3]. In comparison, the number of susceptible persons at infinity *S_∞_ >* 0, and can be calculated from the non-linear equation, [5, 6]:

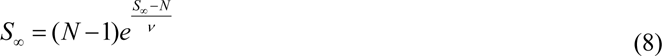

The final number of victims (final accumulated number of cases) can be calculated from:

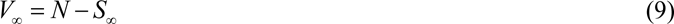

To estimate the duration of an epidemic outbreak, we can use the condition:

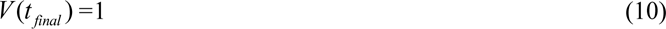

which means that at *t* > *t_final_* less than one person still spread the infection.

### Parameter identification procedure

In the case of a new epidemic, the values of this independent four parameters are unknown and must be identified with the use of limited data sets. A statistical approach was developed in [5] and used in [6-14] to estimate the values of unknown parameters. The registered points for the number of victims *V_j_* corresponding to the moments of time *t_j_* can be used in order to calculate *F*_1_*_j_ = F*_1_(*V_j_*,*N*,*ν*) for every fixed values *N* and *v* with the use of (6) and then to check how the registered points fit the straight line (5). For this purpose the linear regression can be used, e.g., [15], and the optimal straight line, minimizing the sum of squared distances between registered and theoretical points, can be defined. Thus we can find the optimal values of *α*, *t*_0_ and calculate the correlation coefficient *r*.

Then the F-test may be applied to check how the null hypothesis that says that the proposed linear relationship (5) fits the data set. The experimental value of the Fisher function can be calculated with the use of the formula:

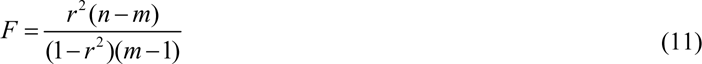

where *n* is the number of observations, *m*=2 is the number of parameters in the regression equation, [15]. The corresponding experimental value *F* has to be compared with the critical value *F_C_* (*k*_1_, *k*_2_) of the Fisher function at a desired significance or confidence level (*k_1_* = *m* −1, *k*_2_ = *n* − *m*), [16]. When the values *n* and *m* are fixed, the maximum of the Fisher function coincides with the maximum of the correlation coefficient. Therefore, to find the optimal values of parameters *N* and *v*, we have to find the maximum of the correlation coefficient. To compare the reliability of different predictions (with different values of *n*) it is useful to use the ratio *F* / *F_C_*(1, *n* − 2) at fixed significance level, [17]. We will use the level 0.001; corresponding values *F_C_*(1, *n* − 2) can be taken from [16]. The most reliable prediction yields the highest *F* / *F_C_* (1, *n* − 2) ratio.

## Results

Usually the number of cases during the initial period of an epidemic outbreak is not reliable. To avoid their influence on the results, only *V_j_* values for the period April 9-29, 2020 (47 ≤*t_j_* ≤67; *n*=21; *F_C_*(1,*n*−2) =15.2; see Table 1) were used to calculate the epidemic characteristics. Since during the quarantine, the international people exchange is quite limited, we can apply the SIR model for every country assuming its parameters to be constant (but different for every country) during the fixed period of time. The results of calculations are shown in Tables 2 and 3. To illustrate the influence of data on the results of SIR simulations, the previous estimation for Germany (prediction 1 calculated with the use of *V_j_* values for the period March 28 – April 10, 2020 (35 ≤ *t_j_* ≤ 48; *n*=14; *F_C_*(1*,n* − 2) =18.6; see [13]) are also presented in Table 2.

**Table 2.**
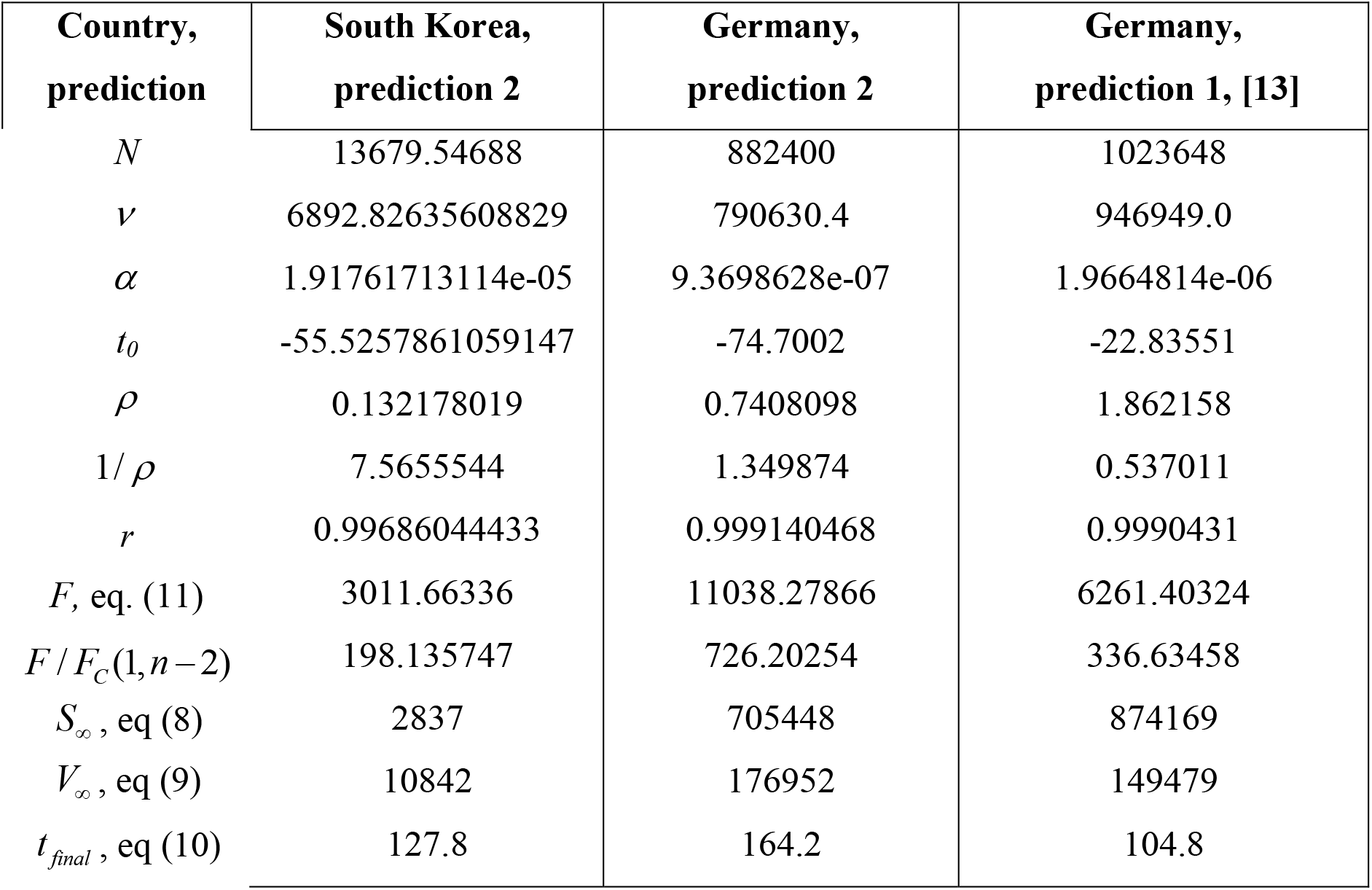
Epidemic characteristics for the Republic of Korea and Germany. Optimal values of parameters, final sizes and days (last two rows).

| Country,<br>prediction | South Korea,<br>prediction 2 | Germany,<br>prediction 2 | Germany,<br>prediction 1, [13] |
| --- | --- | --- | --- |
| $N$ | 13679.54688 | 882400 | 1023648 |
| $\nu$ | 6892.82635608829 | 790630.4 | 946949.0 |
| $\alpha$ | 1.91761713114e-05 | 9.3698628e-07 | 1.9664814e-06 |
| $t_0$ | -55.5257861059147 | -74.7002 | -22.83551 |
| $\rho$ | 0.132178019 | 0.7408098 | 1.862158 |
| $1/\rho$ | 7.5655544 | 1.349874 | 0.537011 |
| $r$ | 0.99686044433 | 0.999140468 | 0.9990431 |
| $F$, eq. (11) | 3011.66336 | 11038.27866 | 6261.40324 |
| $F / F_C(1, n-2)$ | 198.135747 | 726.20254 | 336.63458 |
| $S_\infty$, eq (8) | 2837 | 705448 | 874169 |
| $V_\infty$, eq (9) | 10842 | 176952 | 149479 |
| $t_{final}$, eq (10) | 127.8 | 164.2 | 104.8 |

**Table 3.** Epidemic characteristics for UK, USA and world. Optimal values of parameters, final sizes and days (last two rows).

| Country | UK | USA | World |
| --- | --- | --- | --- |
| $N$ | 479782.4 | 7082400 | 6637317.12000000 |
| $\nu$ | 361758.017613005 | 6312894.63367680 | 3537150.79869746 |
| $\alpha$ | 9.1371956639e-07 | 1.08933738707e-07 | 2.34901375364096e-08 |
| $t_0$ | -47.0679605620665 | -85.9627117094321 | -144.265486657790 |
| $\rho$ | 0.330545378991741 | 0.687687214511608 | 0.0830881587484244 |
| $1/\rho$ | 3.02530322175518 | 1.45414947216984 | 12.0354093178044 |
| $r$ | 0.999363727388945 | 0.999378778826037 | 0.999861835424859 |
| $F$, eq. (11) | 14916.4586251212 | 15278.2115499352 | 68744.3303788117 |
| $F / F_C(1, n-2)$ | 981.345962179029 | 1005.14549670626 | 4522.65331439551 |
| $S_\infty$, eq. (8) | 264883 | 5601247 | 1595881 |
| $V_\infty$, eq. (9) | 214899 | 1481153 | 5041436 |
| $t_{final}$, eq. (10) | 180.1 | 213.5 | 393 |

It can be seen that the previous prediction for Germany (see [13]) were more optimistic. Fresh data sets has showed that the final number of cases in this country could reach 177,000 and their appearance can stop only after August 4, 2020 (see Table 2, prediction 2). The presented second prediction for South Korea is also more pessimistic in comparison with the first one, [8]. In particular, the epidemic stop is expected after June 29, 2020 (see Table 2). These estimations are valid only when the quarantine measures, isolation rate and the coronavisus activity will be same as for the period taken for calculations.

Tables 2 and 3 illustrate that real epidemic outbreaks in Germany, USA and the Republic of Korea probably occurred in November-December 2019, in UK – in early 2020. The real beginning of the global COVID-19 pandemic can be attributed to the beginning of October 2019. It happened in China, most likely in Wuhan, the epicenter of its visible course. Unfortunately, official data from China are very contradictory, which makes their analysis impossible using the SIR model. In any case, the estimations of the *t*_0_ values presented in [6, 7] are no longer relevant. The rather long duration of the pandemic is expected. The last cases could stop to appear only in March 2021 after exceeding the value 5 millions. This long-term prediction is very preliminary, corresponds to the current situation and does not take into account the repeating outbreaks that are possible and are already happening in many countries.

The results about the epidemic hidden periods (time between *t*_0_ and the day when the first COVID-19 case was confirmed), epidemic durations *t_final_* −*t*_0_ and the final numbers of cases *V_∞_* (final sizes) for different countries are presented in Table 4. For the world data, December 8, 2019 was taken as the day of the first laboratory confirmed case in Wuhan, China (according to [18]). The results of previous calculations for Austria, Italy, Spain, France, Moldova, Ukraine and Kyiv from [11, 13, 14] are also shown in Table 4. It can be seen that the longest epidemic durations are expected in the countries with the longest hidden periods (USA, Italy, Germany). Probably, the zero hidden period in France indicates the need of recalculations after obtaining more recent data on the number of cases. The predicted saturation value 129823 for this country (see Table 4) is 4.5% lower than real number of cases 135980, registered on May 7 after 20 days of observations (after April 18). The real numbers of cases in Spain, Moldova and Austria are also higher than predicted saturation levels shown in Table 4.

**Table 4.**
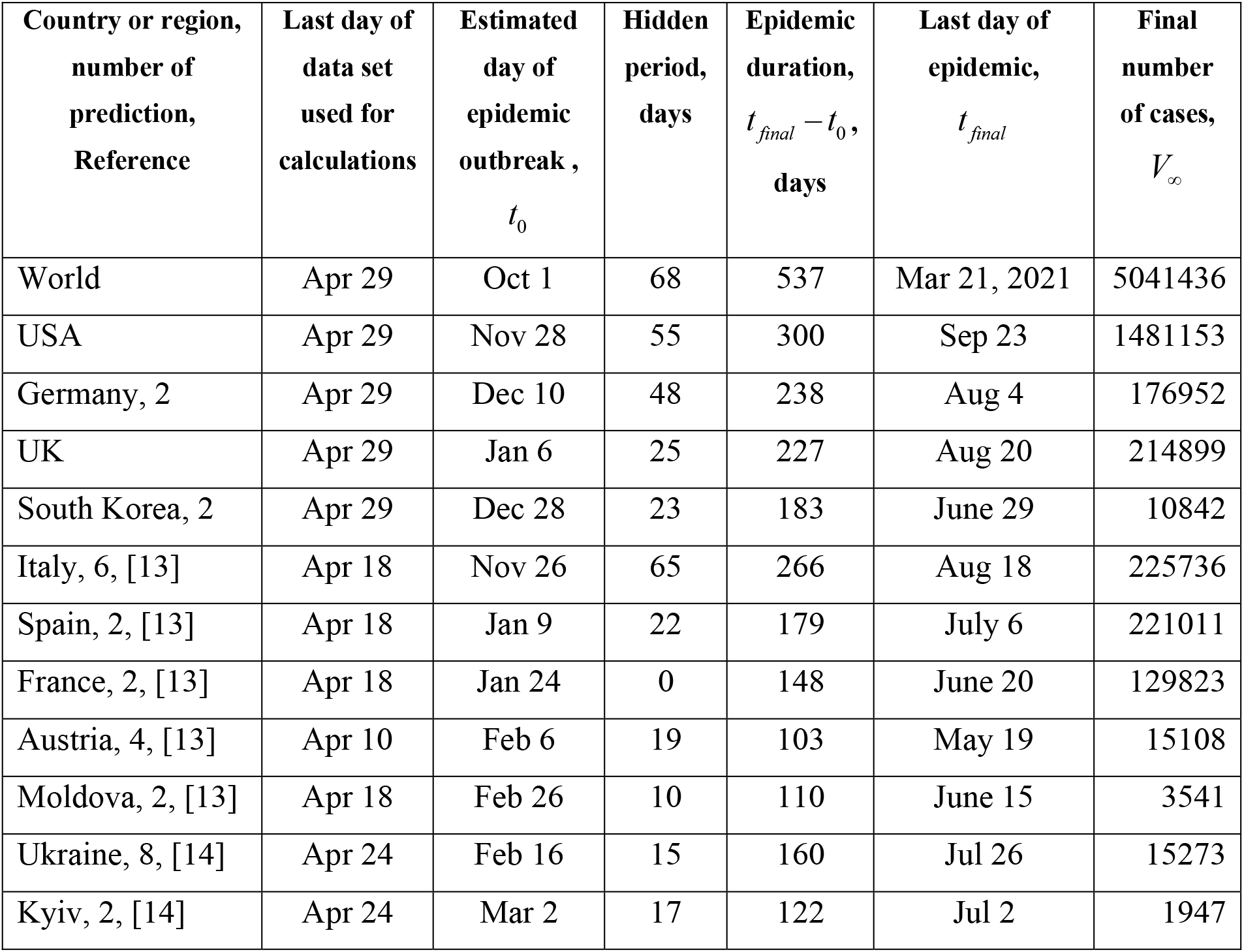
Epidemic characteristics for different countries and regions. Estimated days of epidemic outbreak, hidden periods, epidemic durations, the final days and number of cases.

The SIR curves and markers representing the *V_j_* values taken for calculations (“circles”), comparisons (“triangles”) and verifications of calculations (“stars”) are shown in Figs. 1 and 2 by different colors corresponding to the country or region (World – brown; USA –blue; Italy –green; Spain –yellow; UK – red and South Korea –magenta). Solid lines represent the number of cases (victims) *V*(*t)=I*(*t)+R*(*t*), dashed lines show the number of infected persons spreading the pathogen *I*(*t*). The number of laboratory confirmed cases in Wuhan, China are shown in Fig. 1 by brown “squares”. These values was calculated in [7] with the use of information from [18].

**Fig. 1.**
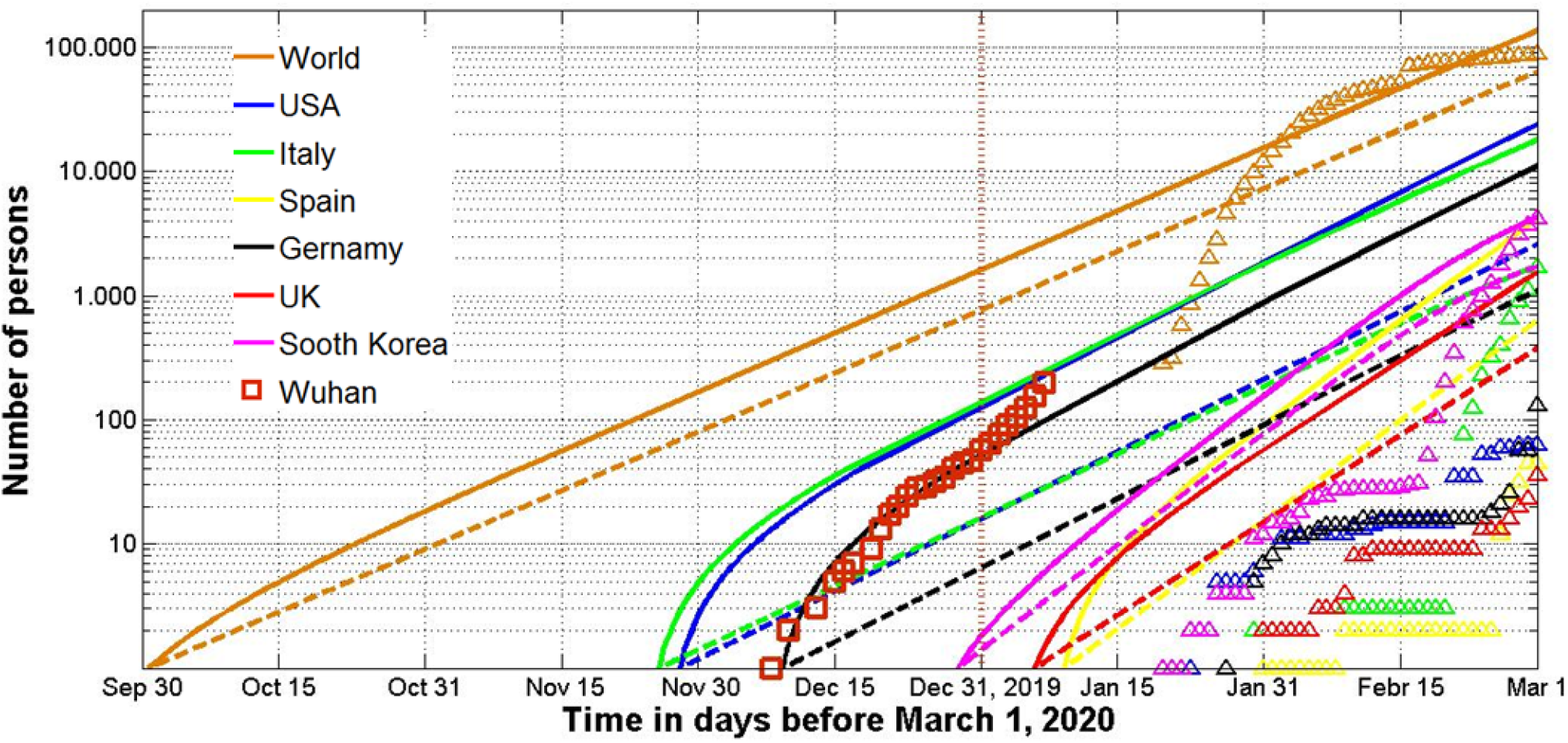
SIR curves (lines) and accumulated number of cases (markers) versus time before March 1, 2020. Numbers of infected and spreading *I* (dashed) and victims *V=I+R* (solid).

**Fig. 2.**
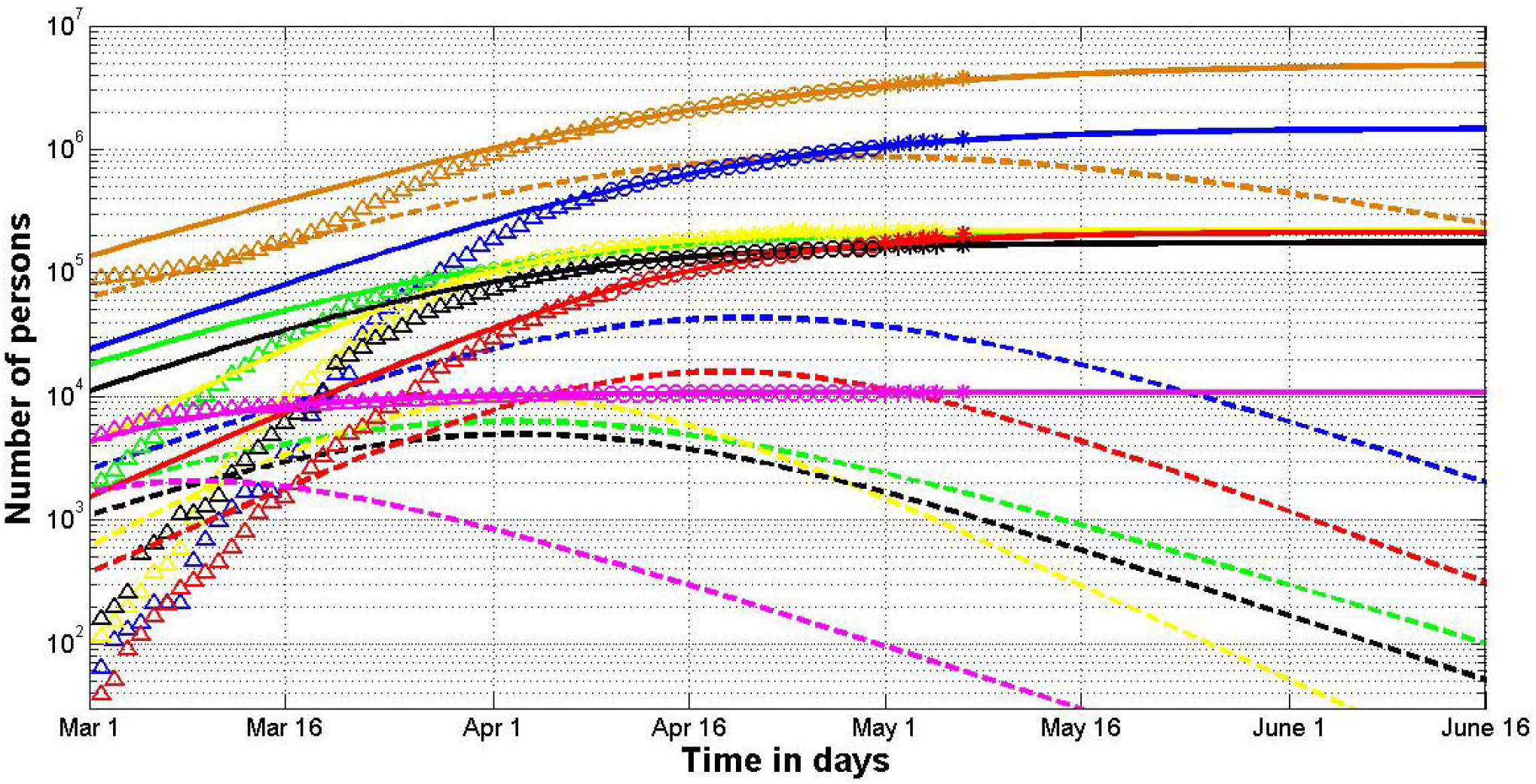
SIR curves (lines) and accumulated number of cases (markers) versus time after March 1, 2020. Numbers of infected and spreading *I* (dashed) and victims *V=I+R* (solid).

Dashed brown curve in Fig. 1 illustrates that more than 600 persons could spread the infection on December 31, 2019 - the day when China notified WHO about the situation in Wuhan (see brown vertical line in Fig. 1). On January 23, 2020 this city was closed but the number of infected persons could be estimated by 3000 with hundreds of cases in USA, Italy and Germany (the official confirmed number of cases was 830 in mainland China, 2 in the Republic of Korea and 1 in USA on this day). The big difference between number of calculated (bold lines in Figs. 1 and 2) and actual cases (“triangles”) is explained by the fact that many infected people do not have symptoms and there were and still are problems with testing. In particular, the hidden periods (time between the estimated epidemic beginning *t*_0_ and the first confirmed case) can be rather long (see Fig. 1 and Table 4).

Recently, there is more and more evidence in the media and literature about the hidden periods of the epidemic. In particular, according to [13] the first COVID-19 could happen in Italy around November 27, 2019 (see Table 4). This results correlates with the information form Giuseppe Remuzzi, director of the Mario Negri Institute for Pharmacological Research that “virus was circulating before we were aware of the outbreak in China”, [19]. Probably the spread of the infection was facilitated by the Military World Games held in Wuhan from 18 to 27 October with the participation of 9,300 athletes from more than 140 countries. Many participants became ill with COVID-19 symptoms and passed the infection on to their families, [20]. A very fundamental statistical analysis [21] showed that the number of cases of pneumonia in the United States in January and February 2020 exceeded last year’s figures and this excess was higher in those states where the actual numbers of reported COVID-19 cases are larger.

## Discussion

### Reliability of predictions

It was already mentioned that during the initial stages of epidemic the registered number of cases is much lower than the real one. This fact reduces the accuracy of any simulation using the registered values. Nevertheless, the prediction have to be performed in order to estimate the final sizes and the durations of epidemics in different countries even with limited accuracy.

Errors caused by incomplete data can be illustrated by two different predictions for the Republic of Korea. Both of them was performed with the use SIR model and the same method of parameter identification. The first one used the data for the period February 17 - March 12, 2020 and predicted: the final accumulated number of cases *V*_∞_ ≈ 8117; *t*_0_ = −9 days and final day of epidemic

March 20, 2020 (see [8]). The results of calculations presented in Table 2, illustrate that the difference in final sizes is about 25%, but the epidemic duration predictions differ more than 5 times. Three different estimations for Italy [10, 11, 13] yielded the variation in the final sizes from 111548 to 225736; the hidden period estimates increased by 66 days with the use of more recent data sets.

Since the presented forecasts for USA and world are very long-term, they must be considered as very preliminary and optimistic. The global pandemics dynamics is very unpredictable, since the situation is very different in different countries. In particular, there is no quarantine in Belarus and the word “coronavirus” is prohibited in Turkmenistan. In any case the global dynamics must be updated with the use of new data sets. Unfortunately, the new estimations will probably be more pessimistic.

SIR model allows calculating the average time of spreading infection 1/ *ρ*. For example, the first prediction for South Korea yielded incredible low value of 4.3 hours, [8]. Table 2 yields more realistic figure - 7.6 days. Probably, such big difference is connected with the huge increase of the hidden period for the second prediction. Similar large differences were also observed in two different forecasts for Ukraine and Kyiv [11, 14]. The use of recent data increases the estimation for 1/ *ρ*. Its real value could be calculated only after epidemic stabilization. When the number of confirmed cases tends to the real one, the accuracy of SIR simulation may be rather high. An example is the latest forecast for Austria [13]. After 28 days of observation the predicted final size (see Table 4) is only 3.6% lower than the real number 15673 (May 7, 2020).

### Control of testing and relaxation of quarantine

SIR simulation could be used to control the situation with COVID-19 testing. For example, in Ukraine the PLC tests were introduced for pneumonia patients and medical staff only after April 10, 2020. To estimate how the change in testing algorithm affected the epidemic dynamics in Ukraine and Kyiv, two series of calculations were performed in [13, 14] with the use of data sets for the periods March 28 – April 10 and April 11-24. Figs. 3 and 4 illustrates the results of these estimations. The corresponding SIR curves (solid for the second data set and dashed for the first one) are very different. Delayed proper testing could cost Ukraine at least 9,000 additional patients and an epidemic duration increase of 47 days (compare blue lines in Fig. 3). That is why the maximum PCR testing (especially for medical staff and patients with pneumonia) can be recommended in all countries as an effective means of reducing the scale and duration of the pandemic.

**Fig. 3.**
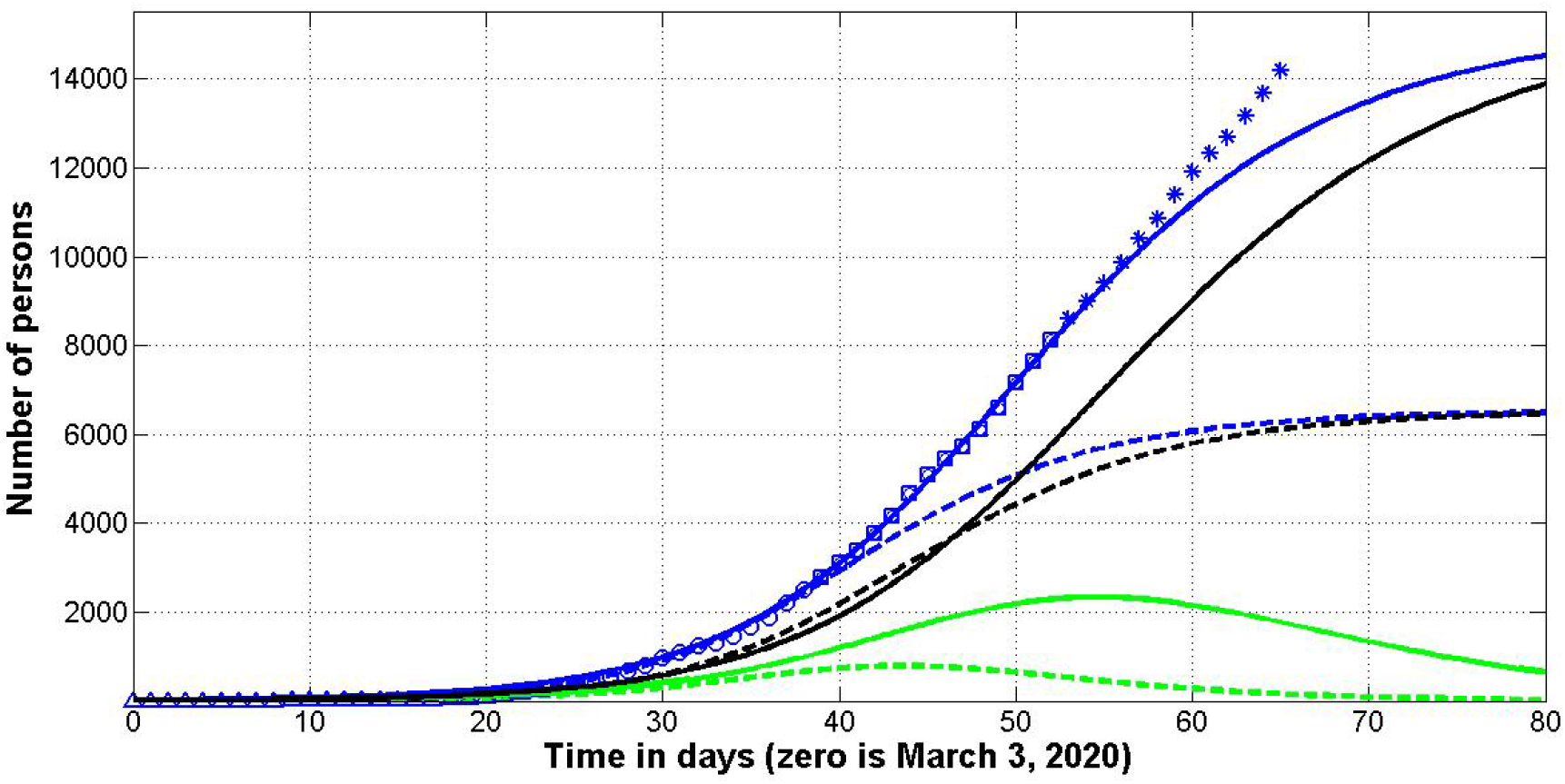
Ukraine: SIR curves (lines) and accumulated number of cases (markers) versus time. Numbers of infected and spreading *I* (green), removed *R* (black) and victims *V=I+R* (blue). Dashed lines correspond to the fist data set (March 28 – April 10) shown by “circles”, solid – to the second data set (April 11-24), shown by “squares”. “Triangles” show the accumulated numbers of cases before March 28, “stars” – after April 10.

**Fig. 4.**
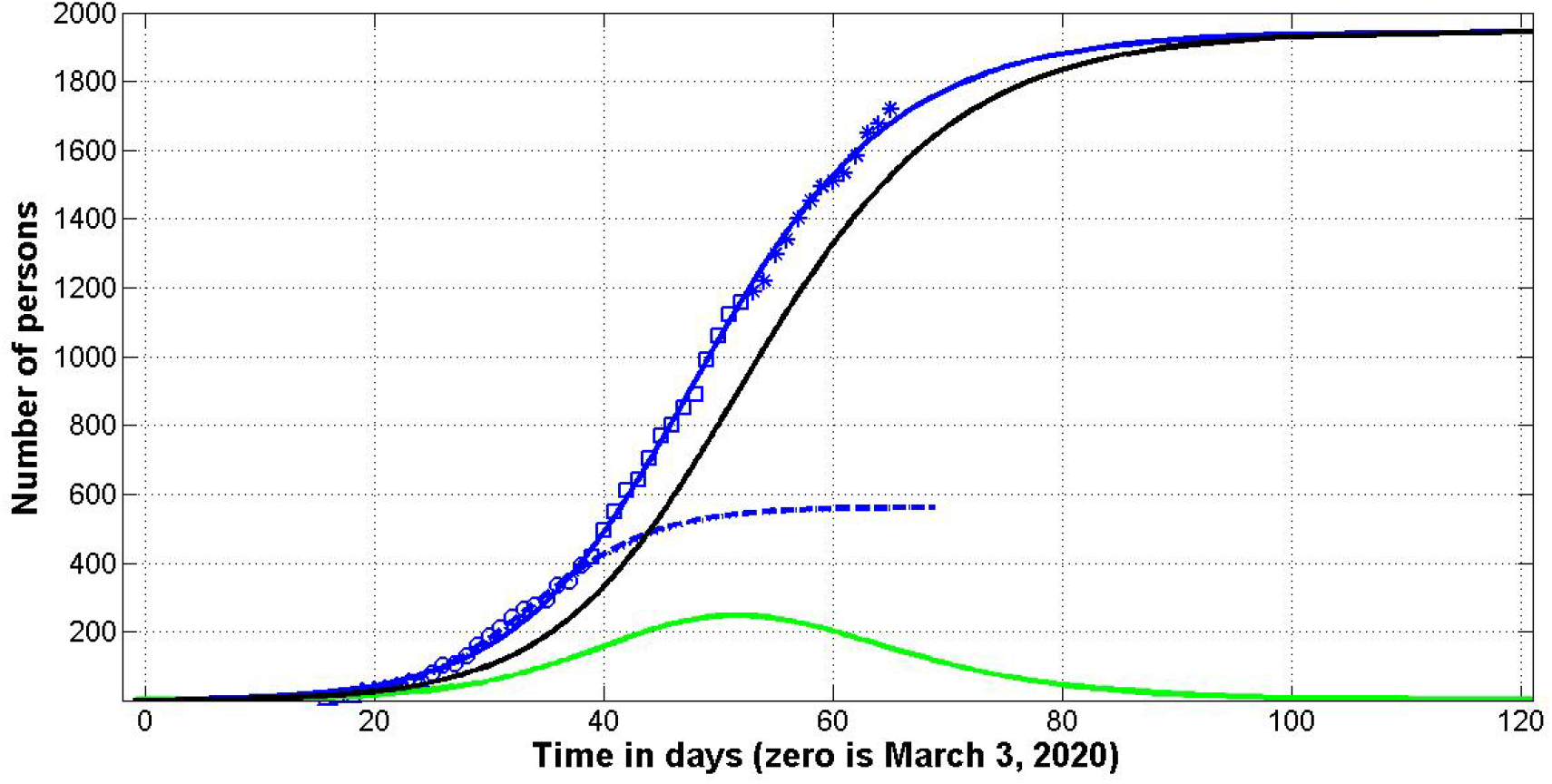
Kyiv: SIR curves (lines) and accumulated number of cases (markers) versus time. Numbers of infected and spreading *I* (green), removed *R* (black) and victims *V=I+R* (blue). Dashed line corresponds to the fist data set (March 28 – April 10) shown by “circles”, solid – to the second data set (April 11-24), shown by “squares”. “Triangles” show the accumulated numbers of cases before March 28, “stars” – after April 10.

The calculated SIR curved were used to control the epidemic dynamics after April 24, 2020 (see “stars” in Figs. 3 and 4). It can be seen, that the situation in Kyiv develops according to the second prediction (see Fig. 4), but the number of cases in Ukraine increases much rapidly in comparison with the second prediction shown in the Fig. 3 by solid blue line. The obvious differences in the epidemic dynamics can be explained by the fact that the situation with testing in the Ukrainian province is much worse than in the capital. It is possible that the dynamics was also affected by the large number of quarantine violations in the province during the orthodox Easter celebrations.

The problem of easing quarantine has become urgent in many countries. The SIR *I*(*t)* curves can be used to estimate possible risks. For example, green line in Fig. 4 illustrates that in Kyiv the number of infected persons (they may feel completely healthy or have mild symptoms) is estimated by 100 on May 11, 2020 (the day of quarantine easing, *t_j_* =68). It means that the probability to meet such person is rather low *p ≈* 3,4 × 10^−5^. But if you have *N* contacts, the probability of meeting at least one infected person increases according to the formula:

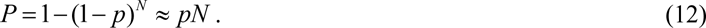

Therefore, a person on duty in the subway or a trolleybus driver is quite likely to meet an infected person during, for example, 10 days of work (until the value of *p* decreases). Meeting an infected person does not mean getting infected, so masked mode and distance in transport and other public places must remain mandatory. Workers in transport, trade, pharmacies, police (all of whom are forced to have many contacts) must be provided with enhanced protection. People at risk should continue to refrain from traveling, visiting indoors and minimizing visits to medical facilities. Where possible, distance work and study should be maintained.

For ordinary Kyiv citizens working in small groups, the risk of meeting an infected person depends on the transport situation. If cars and metro stations, land transport will be regularly decontaminated, the values of *N* and *P* for passengers will be small. Everyone (not only in Kyiv) can assess their own level of risk (allowable probability) using formula (12).

### Could the pandemic be avoided?

The answer to this question may be rather positive. This is evidenced by the experience of Hong Kong, which introduced the relevant measures on December 31, 2019 as soon as alarming information from Wuhan appeared on a social media platform. The Hong Kong authorities immediately closed “transport ties with Wuhan”; increased “vigilance and temperature screenings at every border checkpoint, including the city’s international airport and high-speed railway station in West Kowloon”, the hospitals were told “to report further cases of “pneumonia of unknown origin”, [22]. As a result the accumulated number of cases in Hong Kong was 1045 on May 9, 2020. For example, the number of infected people in Kyiv was 1.7 times higher.

## Conclusions

The SIR (susceptible-infected-removed) model and statistical approach to the parameter identification are able to make some reliable estimations for the epidemic dynamics, e.g., the real time of the outbreak, final size and duration of the epidemic and the number of persons spreading the infection versus time. This information may be useful to regulate the quarantine activities and to predict the medical and economic consequences of the pandemic. The pandemic outbreak probably occurred in China not later than the end of September 2019, it could continue beyond mid-March 2021, and the number of infected people in the world could exceed 5 million.

## Data Availability

data are in manuscript

## Acknowledgements

I would like to express my sincere thanks to Gerhard Demelmair, Ihor Kudybyn, Nina Basiuk and Volodymyr Borysenko for their help in collecting and processing data.

